# Understanding needs and solutions to promote healthy ageing and reduce multimorbidity in Rwanda: A protocol paper

**DOI:** 10.1101/2024.12.02.24318282

**Authors:** Alemayehu Amberbir, Callixte Cyuzuzo, Michael Boah, Francois Uwinkindi, Chester Kalinda, Tsion Yohannes, Sandra Isano, Robert Ojiambo, Carolyn Greig, Justine Davies, Lisa R Hirschhorn

## Abstract

**Background:** Ageing is often accompanied by chronic diseases, multimorbidity, and frailty, increasing the need for clinical and social care to support healthy Ageing and manage these conditions. We are currently in the UN Decade of Ageing and there is a growing focus on the need to prevent or delay some of these conditions through the “Healthy Ageing” initiative of the World Health Organization. However, there are limited data available to inform prioritisation of interventions, particularly for countries in sub-Saharan Africa.

**Methods:** This study has five interlinked work packages (WPs), designed to understand the current needs for older people in Rwanda, health system capacity and possible solutions to unmet need. First, we will conduct a household survey in the City of Kigali (predominantly urban) and Northern Province Burera district (predominantly rural) to determine the burden of multimorbidity, frailty, access to care, and experiences and responsiveness of care in older people. This work will be supplemented by secondary analysis of data from the Rwandan STEPwise approach to non-communicable disease (NCD) risk factor surveillance (STEPs) survey of 2021. Second, we will conduct a health facility readiness assessment and healthcare provider survey to assess health system capacity and gaps to deliver effective primary care to older people in Rwanda. Third, to capture the voice of older people, we will explore the quality of healthcare as experienced by them using in-depth interviews (IDIs). In Fourth, we will synthesise data using mixed methods to understand barriers to access to quality of care in older age based on a 3-delays framework (seeking, reaching, and receiving quality health care). Finally, the project will culminate in a stakeholder workshop to ensure results are contextually appropriate and disseminated, and gaps identified are prioritized to design novel interventions to promote healthy ageing in Rwanda and the region.

**Discussion:** This study will deliver impactful research by using multiple methodologies and working with in-country partners to develop a deep knowledge and understanding of health care systems experienced by older people in Rwanda. It will also provide a framework for sustainable healthy ageing research and policy engagement to benefit older adults living in Rwanda and inform similar work in Low- and Middle-Income countries (LMICs) during this Decade of Healthy Ageing and beyond.

**Strengths and limitations of this study:**

- Strengths of this study include our cross-disciplinary mixed methods health systems research, implementation research and a large population-based survey design;
- Our population-based study will cover over 127 villages in the city of Kigali (predominantly urban) and Burera district in Northern part of Rwanda (predominantly rural) covering a total sample size of 4280 increasing external validity of the study;
- It will provide, to our knowledge for the first time, a picture of public-sector health facility care seeking behaviors and health utilization experiences among older individuals in urban and rural Rwanda;
- Further strengths include our stakeholders’ involvement and engagement which will lead to strategies for translation of the findings in to practice and impact;
- Limitations of this study will include the cross-sectional nature of the study limiting our ability to assess a cause-and-effect relationship. Moreover, some of the chronic conditions (heart and respiratory disease, high cholesterol) and HIV in the study will rely on self-report of a diagnosis;
- Our study is further limited in that we will not collect all dimensions of the health systems responsiveness domains. Moreover, the study will not be representative of health centers and health care workers in private or higher-level health care facilities in Rwanda.

## Background

Despite the growing global population of older people and recognition of their increased health and social care needs, this group has been largely neglected in global development goals.^1^ In redressing this gap, the World Health Organization (WHO) has launched the ‘Decade of Healthy Ageing’ 2021-2030 to improve the lives of older people and those who look after them.^2^ Prolonging the human lifespan is a remarkable achievement, but it poses substantial challenges. The increase in chronic diseases, multimorbidity, and frailty that accompany ageing are especially challenging in low- or middle-income countries^3^ – where a growing number of people aged over 60 years live and where background poverty, rapidly changing societies, and under-resourced health systems to place as yet unknown demands on individuals and health care services. The WHO sets out an urgent need for multisectoral collaboration including government, civil society, and academia to address the needs related to this changing demographic and to enable older adults to live with dignity in a healthy environment.^2^ However, 75% of all countries in the world – including Rwanda - have no or limited data to allow effective plans to improve healthy ageing.^2,4^ In 2021, Rwanda developed a National Older Persons Policy including priority health interventions but its implementation remains very limited.^5^

Emerging evidence suggests that the burden of multimorbidity is especially high among older people in Low-and- middle income countries (LMICs).^3,6^ Additionally, the prevalence of age-related frailty (a condition strongly associated with higher rates of death, and hospitalization) seems to be higher at lower ages in LMICs compared with high income countries (HICs).^4^ Frailty adversely impacts on most characteristics identified by WHO as necessary for healthy ageing.^7,8^ Despite this growing burden, studies to date indicate that health care systems, including primary care, are ill-suited to provide coordinated preventive, curative, and rehabilitative care for the ageing population.^9,10^

There is also little knowledge on the perceptions of ageing and needs and priorities of older people or the people who care for them from LMIC settings. Studies that have been done are limited in scope.^11^ Knowledge of ageing in societal contexts is crucial for achieving improvements in health and realizing the economic benefits that older people bring.^20^ While Rwanda has extended Universal Health Coverage (UHC) through the innovative Community-Based Health Insurance (CBHI) programme, this is not necessarily responsive to the needs of older people; for example, there are limited medical personnel trained in geriatrics and gerontology.^12^ The results of this cross-disciplinary study will therefore provide evidence to design novel interventions to maintain health ageing and inform policy and planning.

## Methods

The research will achieve our goals through the five interconnected objectives corresponding to five work packages (WPs) designed to provide a comprehensive mapping of existing burdens of age-related and other comorbidities and the capacity of the health system to meet these needs. This understanding from multiple perspectives is needed to understand where capacity is needed to address structural and societal barriers to effective health care for ageing individuals in Rwanda. Our longer-term goal is to develop future studies to test prioritized, co-developed interventions to strengthen care and health among this population and create a road map for Rwanda and other countries in the region to meet the goals of the UN Decade of Healthy Ageing 2021-2030.

Our objectives are:

1. To assess the local prevalence of multimorbidities (frailty, cognitive function, mobility, chronic conditions), patient-reported outcomes (disability, quality of life) and responsiveness of healthcare to needs through using both primary and secondary household survey dataset;
2. To assess the readiness of primary health care sites in rural and urban Rwanda, capturing health care provider’s knowledge, attitude, and practice towards ageing and related comorbidity;
3. To explore the quality of healthcare as experienced by older people randomly selected from the household survey in rural and urban Rwanda;
4. To utilise the three-delays model and data from objectives 1-3 to illustrate barriers to access to quality of care;
5. To conduct stakeholder workshop to ensure results are contextually appropriate and disseminated, and interventions are prioritized for co-development and testing.

### WP 1a: Quantitatively analyze the burden of multimorbidity, frailty, access to care, and experiences and responsiveness of care in older people

This WP has two main areas of research: primary household survey and analysis of existing nationwide secondary dataset on chronic disease morbidity.

First, we will analyse the recent WHO National NCDs risk factors STEPs survey (conducted from November 2021 to January 2022) to understand the current burden of multimorbidity and risk factors among older people in Rwanda, their patterns of access to care, and unmet need. Using the previous rounds of the WHO STEPwise survey conducted in Rwanda,^13^ the study will quantify the changes (if any) in the prevalence of NCDs by decomposition of the risk factors into compositional characteristics and behavioural (coefficients) factors. Furthermore, the study will assess the contribution of various risk factors to changes in the prevalence of NCDs.

#### Description of the dataset

The data-owners are the Rwanda Biomedical Centre (RBC) and Ministry of Health, who are partners on this study. The WHO STEPs data^14^ provides a nationally representative snapshot of prevalence of selected chronic disease conditions and access to care for older people in Rwanda. In brief, the study was a population-based survey of adults aged 18-69 years conducted nationally using the standard STEPs methodology.^14^ A total of 5,676 adults included in the study using a multi-stage cluster sampling technique. The modules used in the survey included cardiovascular disease (CVD), diabetes, cervical cancer, tobacco use, alcohol use, diet, physical activity, road traffic accidents, violence, and oral health. Variables on whether conditions are diagnosed and treated allow the study of care access. Additionally, data were collected on demographics and socio-economic circumstances.

#### Data analysis

The independent variables in this research project are socio-demographic characteristics (age and gender); behavioral information characteristics (tobacco smoking and alcohol consumption, physical inactivity and diet); biochemical measurements (blood sugar and cholesterol) and physical measurements (height, weight, waist circumference and blood pressure). The dependent variables are blood pressure, blood sugar and multimorbidity, where their presence or absence will be evaluated. During the analysis, both multivariate decomposition approach and multilevel mixed model will be employed to analyze the data. For access to care analyses, cascade of care approach will be used. We will construct cascade from these variables on whether participants had been previously tested, diagnosed, treated, or controlled for diabetes and hypertension. Gender, location, and wealth will be used to consider effects of equity on outcomes.

### WP 1b: Conduct a household survey in rural and urban areas to assess prevalence of morbidities (cognitive function, and anxiety and depression), frailty, disability, quality of life), and experiences of healthcare accessed

#### Study settings

This household survey will be conducted in the City of Kigali which includes predominantly urban districts (Gasabo, Kicukiro and Nyarugenge) and a predominantly rural Burera district in the Northern part of Rwanda to understand similarities and differences in study outcomes. Rwanda is a low-income country. Approximately 10% of the population are over 50 years old, retirement age and life expectancy are 65 and 69 years, respectively. The old-age dependency ratio (the number of people age 65 years per 100 people 15-64 years) is high at 5.3.^12^

#### Participants

Similar to the approach taken in other studies where population life expectancy is low,^3,10^ older people are defined as over 40 years old for the purpose of this study. Previous studies in Burkina Faso^3,10^ and South Africa^15^ have also demonstrated levels of frailty and multimorbidity in people over 40 at much higher levels than one would expect in a high-income settings population of the same age.

#### Sample size and sampling techniques

To detect a frailty prevalence of 6% (conservative, using the Fried frailty score)^7,8^ with a margin of error of +/-0.5% with 95% confidence would require a sample of 3852 individuals. Accounting for non-response of 10%. we will recruit 4280 individuals. The sampling frame to be used for the household survey will be the updated frame from the recent 2022 Population and Housing Census conducted in Rwanda. We will utilise stratified two-stage cluster sampling technique designed to yield representative results for the selected urban and rural areas for the indicators to be measured. In the first stage, 127 target villages (enumeration areas) will be selected from the sampling frame using a probability proportional to size strategy for urban and rural areas in the four districts (Gasabo, Kicukiro, Nyarugenge and Burera). The number is chosen on the assumption that, given the distribution of the population aged 40 years or older in the selected districts, it will be adequate to obtain the anticipated sample size. The total number of enumeration areas (EAs) will be allocated proportionally to the selected districts to determine the number of EAs to be selected per district.

In the second stage, after the selection of the EAs, individuals aged 40 years or older will be generated with assistance from the National Institute of Statistics of Rwanda (NISR) in collaboration with village community health workers. According to information from the NISR, it is presumed that in every EA, there will be at least thirty households with an individual within the target age group. As a result, a minimum of thirty households will be selected in each of the EAs. Only one member will be interviewed per household. The details of the number of EAs and individuals to be sampled are as follows:

**Table 1:**
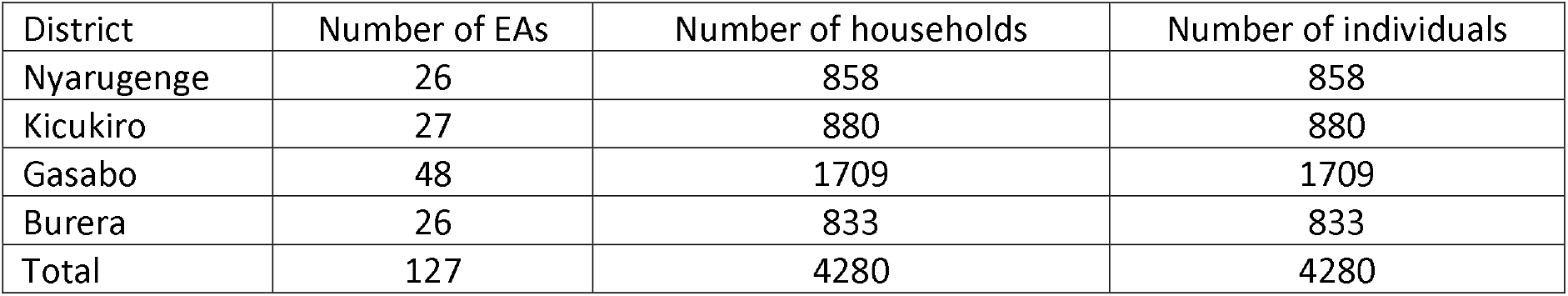
Distribution of the number of households and individuals to be sampled according to the study districts in Rwanda.

| District | Number of EAs | Number of households | Number of individuals |
| --- | --- | --- | --- |
| Nyarugenge | 26 | 858 | 858 |
| Kicukiro | 27 | 880 | 880 |
| Gasabo | 48 | 1709 | 1709 |
| Burera | 26 | 833 | 833 |
| Total | 127 | 4280 | 4280 |

#### Data collection and description of variables

The survey will be carried out by trained data collectors at participants’ homes or other convenient places within study area. Data will be entered onto electronic tablets. We will use a questionnaire previously used in South Africa and Burkina Faso ^9,10^ to collect demographics (age, sex, education level, area of residence); socio-economics (current employment, whether in receipt of health insurance, type of insurance, household income, yearly household expenditure, household assets); carer roles (whether the respondent provides care for others, and who, and how often); medical history and medication use (self-reported); healthcare seeking in previous 3 and 12 months (if so, what type of facility, and if not, why not);^16^ disability;^17^ social networks and social support;^18,19^ anxiety and depression;^20^ cognition;^20^ Quality of Life (QoL) (EQ-5D-5L);^21^ met need for care (and by whom need is met);^15^ and responsiveness of healthcare sought;^16^ We will take measurements of: anthropometry; blood pressure; the short physical performance battery;^22^ and grip strength. The survey will be pilot tested, translated and back translated into Kinyarwanda and cognitive debrief for cultural adaptation.

#### Measures and procedures

Both physical measurements and biomedical assessments will be carried out in the study using validated tools and standardized procedures. Anthropometry data from respondents, including weight, height, and waist circumference, will be collected. Data collectors will request participants to remove shoes and heavy clothing before taking their height and weight measurements. The weight in kilogrammes will be measured once and to the nearest 0.1 kg using the Omron HN289 digital personal weight measuring scale. A tape measure will be used to measure participants’ height and waist circumference. Participants’ height will be measured twice for a consistent reading, but only one reading will be captured in the REDCap database. A third measurement will be taken in instances where the first two readings diverge, and an average of the three readings will be taken. The waist circumference will be measured at the umbilicus level after deep expiration while the respondent is in a standing position.

The Omron HEM-7322 automatic digital blood pressure measuring machine (OMRON Healthcare Co., Ltd, Kyoto, JAPAN) will be used for measuring blood pressure. Three blood pressure measurements will be taken at a five-minute interval in a seated position on the left arm of the respondents. The first measurement will be taken after at least 15-minute rest. We will consider an average of the last two readings. For estimation of random blood sugar, the Accu-Chek active blood glucose glucometer point of care kit, designed for capillary blood glucose testing will be used.^23^

Hand grip strength will be measured using the CAMRY hand dynamometer (CAMRY EH101, Sensun Weighing Apparatus Group Ltd, Guangdong, China).^22^ We will assess both hands three times starting with the dominant hand. Respondents will be asked to squeeze the handle of the dynamometer with maximal effort for at least 5 seconds. The HGS values will be measured in kilogrammes to the nearest 0.1kg. A break of at least 30 seconds will be taken between the readings to prevent fatigue effects.

The Short Physical Performance Battery (SPPB) tool will be used to evaluate the physical performance of respondents. Three domains of the SPPB, namely balance test, chair rise, and walk speed will be assessed in the study.^24^ Balance will be assessed by the respondent’s ability to stand upright in three different positions for 10 seconds each: feet together (side-by-side stand); with one foot partially forward (semi-tandem stand) and with one foot forward (tandem stand).^25^ The chair rise requires respondents to first stand from an armless chair without using their arms for assistance. Then they will be asked to make five rises from the same armless chair without using their arms to assist. The time taken to make the five rises will be recorded using a digital stopwatch on the data collectors’ phones. Walk speed will be measured over a 4-metre course marked out on level ground. Participants will be asked to walk at their regular pace from a standing start. The time taken for participants to complete the walk in each direction will be recorded to the nearest tenth of a second using a digital stopwatch on the data collectors’ phones.

#### Data analysis

Variables will be described using appropriate summaries, depending on distribution and variable type. Outcomes will be summarised with 95% confidence intervals. Where a standard instrument is used, derivation of results will be based on the methods described for the tool. Fried frailty score will be derived from variables collected in the questionnaire and clinical measurement.^7,8^ All results will be presented to assess disparities, by sex, wealth, and location (rural versus urban)). Data on healthcare use (whether the participant had used services, and if there has been none, why not), will be combined with responses on past medical history to ascertain who is seeking care amongst those with known conditions, and for those with known conditions who are not seeking care, what the barriers to care seeking are.

### WP2: Conduct a health facility and health provider survey to measure the health system readiness to deliver effective primary care to older people in Rwanda

#### Study setting

We will conduct a health facility survey to understand the health system’s readiness to deliver effective care to older people. This will include data capture on providers’ perspectives and attitudes towards ageing and related comorbidity. We will include primary health facilities from the City of Kigali (urban site) and Burera district (rural site).

#### Participants

Participants for the survey will be health care providers working in relevant clinical areas at each study facility. These will include hospital management staff, physicians, and nurses. Relevant clinical areas will include outpatient departments and wards.

#### Sample size

Randomly selected primary public/government facilities in the urban and rural districts and healthcare providers working at these facilities in the district will be invited to participate in the survey. We aim to include up to 12 primary healthcare facilities in Burera district and 12 primary healthcare facilities in the City of Kigali. We will include providers in selected primary health care facilities.

#### Data collection

Building on an existing health facility survey developed by the University of Lagos,^26^ we will assess readiness to deliver preventive, diagnostic, curative and rehabilitative care focusing on primary care facilities and community health worker system. Areas will include staffing, supplies, training and experience, structure and availability of key services.

#### Data analysis

Data on health facility and health providers’ survey will be summarised using descriptive statistics and utilising appropriate frameworks.

### WP3: Conduct In-depth Interviews (IDIs) to qualitatively explore the quality of healthcare as experienced by older people and providers

#### Study design

A phenomenological qualitative study will be used due to its suitability to explore the lived experience of older people who need health and social care services. This will explore participants’ interpretation of social and health care, their feelings, perceptions, and experiences in seeking these services, and accessing quality care.

#### Participants

Up to 12 IDI will be done in each rural (Burera) and urban area (City of Kigali), bringing the total to 24 IDIs. In-depth interview participants will include older people (age 40 and older) in each rural and urban areas stratified by gender. Older people who require care (who have and have not sought healthcare in the past 12 months, randomly selected from household survey respondents) will be invited for IDIs. Participants will be asked about their needs for healthy ageing and asked to discuss barriers to seeking, reaching, and receiving quality health and social care to maintain health and well-being in older age.

#### Sample size

Sample sizes have been determined based on our experience of numbers needed to meaningfully capture perspectives in different groups.^11^ Up to 12 key informants will be included in each urban and rural site. Where necessary, numbers will be increased if saturation has not been achieved.^27^

#### Data analysis

IDIs will be audio recorded, transcribed, translated, and analysed using thematic analysis and a three delay’s framework. A three delays framework will analyze how long it takes participants to seek care and factors that influence their decisions, delays in reaching health and social care providers and the challenges they face, and evaluating delays in accessing quality care and the factors that impact timely and effective quality care delivery.

### WP4: Develop a three delays model utilizing data from objectives 1-3 to illustrate barriers to access to quality of care

To understand common barriers and facilitators to access health and social care to achieve health and well-being in older age based on a 3-delays approach (showing seeking, reaching, and receiving quality health and social care)^28^, we will synthesise data from objectives 1-3 using a convergent parallel mixed methods approach. In developing a matrix showing where barriers or facilitators are experienced across multiple methods and participants, we will show which are likely to be important to overcome or invest in to improve healthy ageing.

### WP5: Conduct stakeholder workshop to ensure results are contextually appropriate and disseminated and prioritize interventions for further co-development and testing

#### Participants

Stakeholder workshop participants will be drawn from the health care, policy, researchers, implementers at community level and other groups working to meet the needs of older population.

#### Sample size

We will purposely select 20 stakeholders; with 5-8 representatives of each type. We have found these numbers to work well in previous settings.^29^

#### Data collection

The workshop will be expert-facilitated 1-day stakeholder workshop with the above groups, using a modified nominal-group technique and voting to achieve consensus.

#### Data analysis

Consensus outputs of priorities will be noted. Workshops will also be audio recorded and analysed thematically using rapid qualitative analysis approach. Analysis will use relevant frameworks and emerging themes.

#### Quality control measures

Quality control procedures will include the training of data collectors, after they are recruited through a rigorous recruitment process. The research team will undertake regular field supervision of enumerators as well as a daily review of collected household and facility-level data to identify discrepancies. We will generate a comprehensive summary report at the end of each week providing an overview of the collected data and highlighting areas including missing data. Measuring instruments such as weighing scales, BP machines, glucometers, and hand dynamometers will be calibrated daily following the standard procedures recommended by their manufacturers.

#### Ethics

The study has received ethics approval from the Rwanda National Ethics Committee (RNEC) (Reference no: RNEC262/2023), Northwestern University, USA (IRB ID: STU00220814) and University of Birmingham, UK (IRB ID: ERN-23-0421). Written informed consent will be sought from all research participants. Most of the research involves answering survey questions and taking part in discussions. All clinical measures are physical (e.g.: chair stands, walk speed, and grip strength). The finger prick test for the point of care blood glucose measurement may cause a small amount of discomfort. There are no alterations to care because of taking part in the research. The participants identified with elevated blood pressure and blood sugar will be referred to the nearest health facilities for further diagnosis and treatment.

## Discussion

We have designed the study to deliver impactful research through combining methods and engagement working with key stakeholders and policy makers in Rwanda. Our group will draw upon a portfolio of expertise in innovative and established methodologies, building on tools developed by the team - including facility and provider’s surveys,^26^ household surveys^3,10^ and qualitative analysis.^11^ The work will use a range of methodologies including implementation research, health service assessments, and epidemiological data analysis. We will conduct stakeholders’ workshop to foster sustainable involvement and engagement in research translation to policy making and impact. Finally, we will provide a framework for sustainable healthy ageing research and policy engagement to benefit older adults living in Rwanda and informing similar work in LMICs during this Decade of Healthy Ageing and beyond.

## Data Availability

Not applicable - a study protocol manuscript

## Declarations

Consent for publication Not applicable.

## Availability of data and materials

Not applicable

## Competing interests

The authors declare that they have no competing interests.

## Funding

The authors would like to acknowledge the financial support of Robert J. Havey, MD Institute for Global Health at the Northwestern University through The Northwestern Havey Institute for Global Health Global Innovation Challenge award (1001E).

## Authors’ contributions

AA, JD, CG, and LHR designed the project and led the submission of the grant application. CC, MB, FU (WP1a co-lead), CK (WP1a lead), TY (WP3 lead), SI (WP3 co-lead) and RO are participating in study design, data collection procedures and approach to the study. All authors critically reviewed, provided feedback, and approved the protocol paper.

## Notes

### Competing Interest Statement

The authors have declared no competing interest.

### Funding Statement

The study was funded by the Robert J. Havey, MD Institute for Global Health at the Northwestern University through The Northwestern Havey Institute for Global Health Global Innovation Challenge award (1001E).

### Author Declarations

The study has received ethics approval from the Rwanda National Ethics Committee (RNEC) (Reference no: RNEC262/2023), Northwestern University, USA (IRB ID: STU00220814) and University of Birmingham, UK (IRB ID: ERN-23-0421).

